# High-Density Surface Electromyography Detects Fiber-Type-Related Profiles and Disease-Relevant Fatigue Trajectories in Amyotrophic Lateral Sclerosis

**DOI:** 10.64898/2026.10.20.26366045

**Authors:** Ernesto H. Bedoy, Maia Brown, Breanna Sullivan, Michael A. Christofidis, Tawfiq Al-Lahham, Douglas Weber, Christi L. Kolarcik

## Abstract

Fiber-type composition contributes to contractile behavior and fatigue resistance. Characterizing these physiological differences could help study and monitor disease-related changes. Direct fiber-type measurement requires biopsy, whereas animal findings may not fully translate to humans. High-density surface electromyography (HD-sEMG) may offer repeatable, noninvasive assessment. We recorded HD-sEMG from tibialis anterior (TA) and soleus, muscles with contrasting fast- and slow-twitch-like physiology, during sustained contractions at 30% and 50% maximum voluntary contraction in participants with amyotrophic lateral sclerosis (ALS; *n* = 7) and healthy controls (*n* = 10). Given fast-fatigable motor unit vulnerability in ALS models, we compared TA-soleus profiles and fatigue-related changes. Amplitude, spectral, motor unit firing, and motor unit action potential (MUAP) features were quantified in the first 10 s of stable contraction, with firing and MUAP features derived from decomposition. Exploratory fatigue analyses assessed early-to-late changes and trajectories. Early-window root mean square (RMS), spectral power, firing rate, and MUAP amplitude were higher in TA than soleus in both groups. Group x Muscle interactions were nonsignificant. Despite higher early values, TA showed larger fatigue-related declines in high-frequency power, RMS, and firing rate than soleus, particularly in ALS-affected TA. These findings suggest HD-sEMG detects muscle-type-related electrophysiological differences, with ALS-related changes more evident during fatigue.

## 1. INTRODUCTION

Skeletal muscles differ in fiber-type composition, recruitment strategy, contractile speed, metabolic profile, and fatigue resistance (Bottinelli and Reggiani, 2000; Johnson et al., 1973; Schiaffino and Reggiani, 2011). Human biopsies can quantify Type I, IIa, and IIx fibers, while animal studies can link fiber composition to contractile and fatigue behavior (Bloemberg and Quadrilatero, 2012; Bottinelli and Reggiani, 2000; Schiaffino and Reggiani, 2011). However, animal findings may not fully translate to humans, and biopsies are invasive, regionally limited, and impractical for repeated measurements across muscles or disease stages (Elder et al., 1982; Henriksson-Larsén et al., 1983). Noninvasive approaches to characterize functional muscle-type differences could therefore benefit clinical and translational studies.

Surface electromyography (EMG) reflects motor unit activity and fiber action potentials, as well as volume conduction and recording geometry (Merletti and Farina, 2016). In this paper, “fast-twitch-like” and “slow-twitch-like” describe broad physiological characteristics associated with fiber-type composition, contractile behavior, and fatigue resistance rather than directly identified fiber types. Although EMG does not measure fiber type, its amplitude, spectral content, firing behavior, and fatigue-related changes can reflect these physiological differences (Cifrek et al., 2009; Merletti et al., 2005; Merletti and Farina, 2016). High-density surface electromyography (HD-sEMG) uses closely spaced contacts to quantify raw EMG amplitude and spectral features alongside decomposition-derived firing and motor unit action potential (MUAP) morphology (De Luca et al., 2015, 2006; Nawab et al., 2010).

The tibialis anterior (TA) and soleus provide a useful physiological contrast. TA is a phasic dorsiflexor with a relatively higher proportion of Type II fibers, whereas soleus is a postural plantarflexor with predominantly Type I fibers and greater fatigue resistance (Bottinelli and Reggiani, 2000; Johnson et al., 1973). Neither muscle is purely fast- or slow-twitch, and their surface EMG signals also differ because of depth, pennation, electrode placement, and volume conduction (Lowery et al., 2004; Merletti and Farina, 2016; Zwarts and Stegeman, 2003). Thus, TA-soleus HD-sEMG contrasts may indicate muscle-type-related electrophysiological function without directly identifying fiber composition.

ALS may alter these contrasts through denervation, collateral reinnervation, and motor unit remodeling (de Carvalho et al., 2008; Duleep and Shefner, 2013; Henderson and McCombe, 2017; Martineau et al., 2018). Because reinnervation can change the fiber composition and size of surviving motor units, ALS could either mask or exaggerate TA-soleus differences. Rodent ALS models show preferential early vulnerability of fast-fatigable motor units (Frey et al., 2000; Hegedus et al., 2008; Martineau et al., 2018; Pun et al., 2006). Human evidence is more limited. Previous surface EMG studies, including those using HD-sEMG, have identified motor unit abnormalities in ALS, including evidence of motor unit loss, altered discharge behavior, and recruitment-related hyperexcitability (Bashford et al., 2020; Nishikawa et al., 2022; Noto et al., 2023). It remains uncertain whether ALS modifies the TA–soleus electrophysiological profile or the fatigue-related trajectories of these muscles.

The primary objective was to determine whether early-window HD-sEMG amplitude, spectral, firing, and MUAP features distinguish TA from soleus in healthy participants in directions consistent with fast- and slow-twitch-like physiology. We hypothesized that TA would show higher root mean square (RMS) amplitude, low- and high-frequency power, firing rate, and MUAP amplitude than soleus in healthy participants. The secondary objective was to assess whether this early-window profile persists in ALS and whether early-window features differ by group. We expected these differences to persist in ALS despite possible motor unit remodeling. Finally, we explored fatigue-related changes during sustained contractions, predicting greater declines in TA, particularly in ALS. Although preferential loss of fast-fatigable motor units could leave a more fatigue-resistant surviving population, reduced motor unit reserve and increased demands on surviving units might instead contribute to greater fatigue-related declines (Frey et al., 2000; Hegedus et al., 2008; Pun et al., 2006).

## 2. METHODS

### A. Participants

We analyzed HD-sEMG recordings from 17 participants (7 with ALS and 10 healthy controls). Participants with ALS met revised El Escorial diagnostic criteria (Brooks et al., 2000). Controls had no known neurological disorder or musculoskeletal condition that would interfere with lower-limb EMG recordings. All participants provided written informed consent, and the study was approved by the University of Pittsburgh Institutional Review Board (Study Number STUDY22120029). Participant characteristics are provided in **Table 1**.

**Table 1.** Cohort characteristics and ALS clinical descriptors. Part A summarizes the study groups, including age and sex distribution. Part B provides subject-level disease duration and self-reported lower-limb clinical scores for participants with ALS. Clinical scores ranged from 1 (complete paralysis) to 10 (normal strength). **Abbreviations:** ALS, amyotrophic lateral sclerosis; TA, tibialis anterior; SD, standard deviation; N/A, not available.

| A. Cohort summary |  |  |  |
| --- | --- | --- | --- |
| Group | Participants | Age (years), mean<br>± SD | Male, n (%) |
| Healthy controls | 10 | 63.0 ± 15.8 | 6 (60%) |
| ALS | 7 | 63.1 ± 5.1 | 4 (57%) |

**Table 1. Cohort characteristics and ALS clinical descriptors.**
| B. ALS participant clinical characteristics |  |  |
| --- | --- | --- |
| Subject ID | Approximate disease<br>duration (months) | Lower-limb clinical score |
| ALS01 | > 180 | 1.5 |
| ALS02 | ~21 | 10 |
| ALS03 | ~49 | 5 |
| ALS04 | ~4 | 9 |
| ALS05 | ~12.5 | 7 |
| ALS06 | 9 | N/A |
| ALS07 | 43 | 9 |

### B. Experimental Protocol

HD-sEMG was recorded from TA during ankle dorsiflexion and from soleus during plantarflexion (**Figure 1**). Maximum voluntary contraction (MVCs) was calculated as the mean of three trials using a handheld dynamometer (Hoggan MicroFET2, Salt Lake City, UT). Participants then performed sustained contractions at 30% and 50% MVC to compare recruitment and fatigue demands across intensities. Each trial lasted up to five minutes and ended approximately one minute after the participant reported fatigue, producing variable contraction durations. The stable contraction period excluded the initial force ramp and cessation. Its first (Early) and last (Late) 10 s and five equal time-normalized bins were analyzed (**Figure 1**).

**Figure 1.**
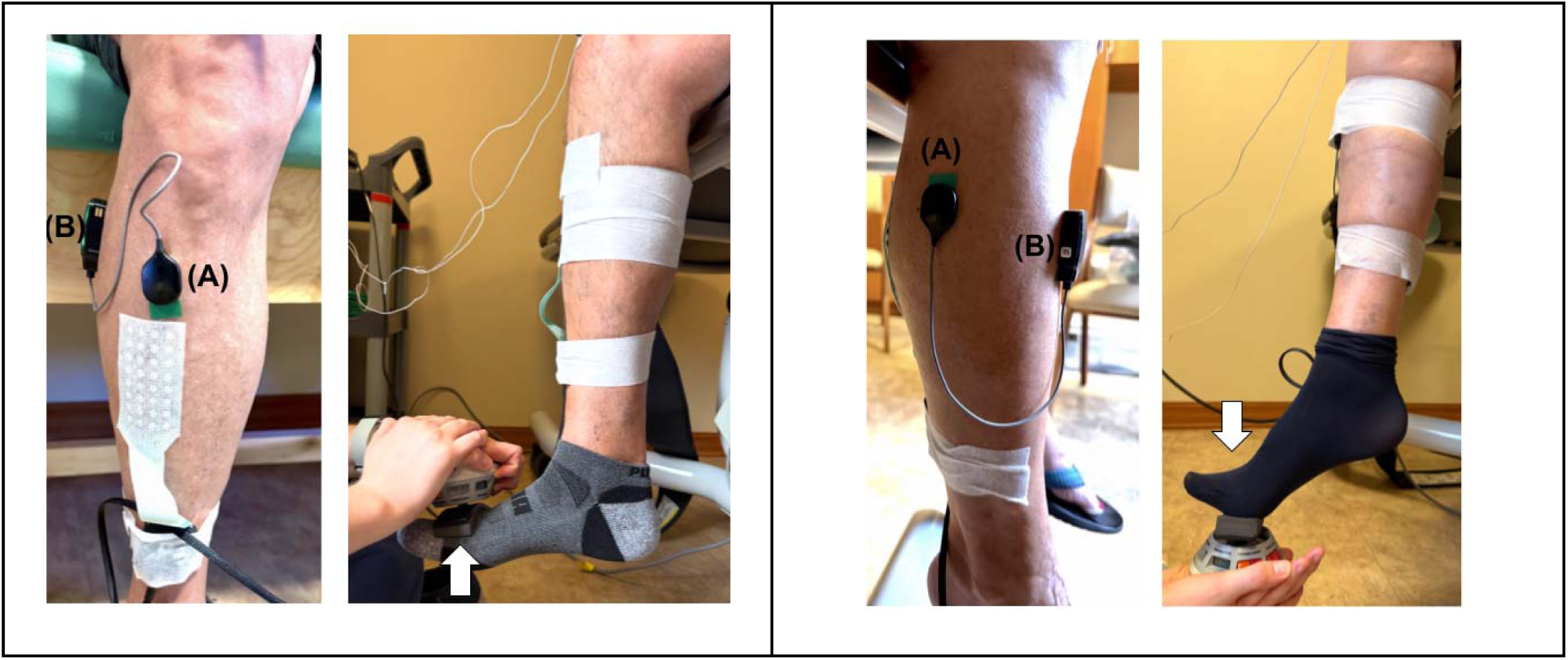
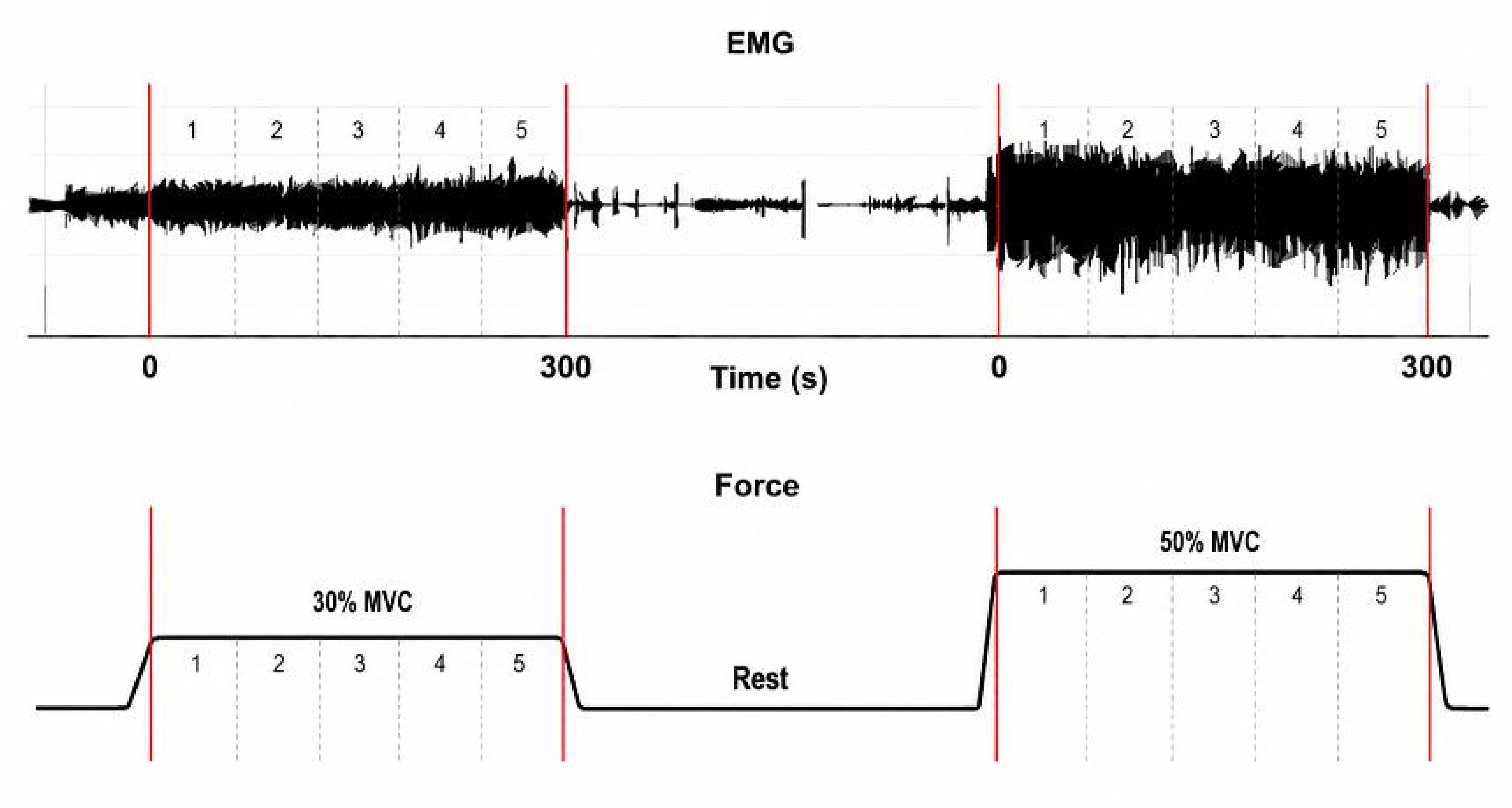
Experimental setup and contraction protocol. HD-sEMG was recorded from tibialis anterior (TA) during dorsiflexion and from soleus during plantarflexion. Representative electrode placements are shown for the HD-sEMG sensor (A) and reference sensor (B). The additional TA grid visible in the setup photograph was not analyzed in the present study. Participants performed MVCs followed by sustained contractions at 30% and 50% MVC. Red lines mark the stable contraction period. Each stable contraction period was divided into five equal time-normalized bins, labeled 1–5, for analysis of fatigue-related changes across the contraction.

This dataset and protocol partially overlap with a previous study (Bedoy et al., 2026), which analyzed spatial and decomposition HD-sEMG from biceps brachii and TA using principal component analysis, with additional exploratory longitudinal and clinical associations. In contrast, the present study uses decomposition-sensor recordings from TA and soleus to examine HD-sEMG profiles and fatigue trajectories associated with their contrasting fast- and slow-twitch-like physiology in healthy participants and those with ALS. Only TA decomposition recordings overlap between two studies.

### C. HD-sEMG Acquisition and Decomposition

Before electrode placement, skin was cleaned and lightly abraded to improve signal quality. HD-sEMG was collected using a Trigno Galileo decomposition sensor (Delsys Inc., Natick, MA, USA), with four circular, 2.5-mm-diameter silver contacts arranged in a 2 x 2 configuration with 5-mm center-to-center spacing. The reference sensor was positioned on soleus for TA recordings and on TA for soleus recordings. Signals were differentially amplified and digitized at 2222 Hz with 16-bit resolution. Raw EMG was high-pass filtered using a fourth-order Butterworth filter with a 20 Hz cutoff. Amplitude and spectral features were derived from filtered EMG, whereas firing and MUAP features were derived from decomposition.

Motor unit spike trains were obtained using NeuroMap v1.2.2 (Delsys Inc.), which implements the Precision Decomposition III algorithm (De Luca et al., 2006; Nawab et al., 2010). Recordings with signal-to-noise ratio (SNR) below 3 dB and motor units with decomposition accuracy below 90%, assessed using Decompose-Synthesize-Decompose-Compare, were excluded. Decompositions with no retained units after quality control were excluded from decomposition-based analyses. Additional acquisition and motor unit quality-control details are provided in the Supplementary Information.

### D. Feature Extraction

We analyzed amplitude, spectral, firing, and MUAP features associated with muscle activity and fatigue (Bashford et al., 2020; Cifrek et al., 2009; Merletti et al., 2005; Merletti and Farina, 2016; Nishikawa et al., 2022; Noto et al., 2023). RMS amplitude quantified overall EMG amplitude. Low-frequency (LF, 20 to 60 Hz) and high-frequency (HF, 170 to 350 Hz) power characterized the lower and higher spectral components, with bands defined a priori. Motor unit firing rate (FR) was obtained from decomposed FR traces, and firing rate variability (FRV) was its coefficient of variation within each analysis window or bin. MUAP amplitude was measured peak-to-peak from spike-triggered averages. MUAP duration spanned the first and last samples where absolute waveform amplitude exceeded 10% of the MUAP’s maximum absolute amplitude. Additional MUAP waveform processing and duration details are provided in the Supplementary Information.

Raw EMG features were calculated by channel and summarized using the median across selected channels. Decomposition features were calculated per motor unit and summarized using the median across retained units. Medians reduced the influence of noisy channels or units.

For Early-to-Late analysis, each feature was extracted from the Early and Late windows (Cifrek et al., 2009; Enoka and Duchateau, 2008; Merletti et al., 2005). Fatigue ratios were calculated as:

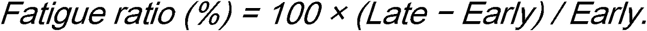

Negative values indicate a decrease while positive values indicate an increase relative to the Early window.

### E. Binned Fatigue Trajectory Analysis

Early-to-Late ratios reflect net change but not its time course. To examine trajectories, features were extracted from five equal time-normalized bins across each stable contraction (Cifrek et al., 2009; Merletti et al., 2005). Bin values were expressed as percentage change from Bin 1 for each subsequent bin, enabling comparison of temporal patterns despite differences in contraction duration. The normalization formula is provided in the Supplementary Information.

### F. Spike Artifact Blanking

To limit transient contamination of binned RMS and spectral estimates, we applied conservative artifact blanking to filtered EMG. Channel-specific thresholds were set at 12 times the robust standard deviation estimate derived from the median absolute deviation of the stable contraction signal. An 8-ms interval on each side of detected artifacts was excluded from raw EMG calculations and MUAP spike-triggered averages. All contractions remained in the primary analysis. Post hoc sensitivity analyses repeated the trajectory analysis after excluding contractions with more than 5% or 10% blanked samples to assess whether recordings with greater artifact contamination influenced the observed patterns. Additional artifact detection, blanking, and sensitivity analysis details are provided in the Supplementary Information.

### G. Statistical Analysis

Linear mixed-effects models were fitted by maximum likelihood, with residual degrees of freedom used for fixed-effects tests. With TA as the reference muscle, Group coefficients represented ALS-versus-Healthy differences within TA. Fixed effects were Group (Healthy vs ALS), Muscle (TA vs soleus), MVC level (30% vs 50%), and the Group × Muscle interaction, with a subject random intercept. The interaction tested whether the TA-soleus contrast differed between groups. RMS, LF and HF power, and MUAP amplitude were log-transformed because of right skew. FR and FRV remained on their original scales. MUAP duration was analyzed descriptively because estimates often approached the 30-ms waveform extraction limit, raising concerns about their reliability for inferential comparisons. Adjusted means of log-transformed features were back-transformed for reporting.

Effect-size analyses quantified TA-minus-soleus contrasts in healthy controls and participants with ALS. We used 2,000 participant-level bootstrap resamples to estimate TA-minus-soleus contrast direction and 95% confidence intervals. For analyses pooled across MVC levels, each participant contributed the mean of their available force-specific paired contrasts. Confidence intervals used the 2.5th and 97.5th bootstrap percentiles (Efron and Tibshirani, 1993), and leave-one-subject-out analyses assessed individual influence. Because soleus recordings were less complete, particularly in ALS (**Table S1**), primary analyses used all eligible Subject × Muscle × MVC observations, with both-muscle and muscle-specific sensitivity checks. For exploratory within-TA comparisons, slopes were averaged across available MVC levels per participant and compared between groups using exact two-sided permutation tests for HF power, RMS, and FR. P-values and bootstrap confidence intervals were reported without adjustment for multiple comparisons. Feature-level findings were considered exploratory. Statistical significance was defined as p < 0.05.

All custom signal processing, feature extraction, and statistical analyses were performed using MATLAB (MathWorks, Natick, MA, USA).

## 3. RESULTS

### A. Cohort Characteristics and Data Availability

Seventeen participants contributed eligible data (7 ALS, 10 healthy controls), with comparable age and sex distributions (**Table 1**). ALS disease duration ranged from approximately 4 months to more than 15 years, and self-reported lower-limb scores ranged from 1.5 to 10 on a 1-10 scale (1 = complete paralysis; 10 = normal strength). The final dataset contained 51 Subject × Muscle × MVC observations. TA recordings were available from 16 participants (9 controls, 7 ALS), and soleus recordings from 12 (7 controls, 5 ALS). Eleven participants (6 controls, 5 ALS) contributed both muscles. Soleus data were less complete, particularly for ALS at 30% MVC (**Table S1**). Primary models included all available observations.

### B. Healthy TA and Soleus Show Distinct Early-Window HD-sEMG Profiles

In healthy participants, TA showed higher early-window RMS amplitude, LF and HF power, FR, and MUAP amplitude than soleus (**Figure 2**; **Table 2**). All five differences were statistically significant (unadjusted p ≤ 0.0011), with TA-to-soleus ratios ranging from 1.95 for MUAP amplitude to 14.55 for LF power. FRV was significantly lower in TA (p = 0.022). TA exceeded soleus at both force levels for RMS, LF and HF power, and FR. MUAP amplitude differences were less consistent across force levels but remained positive when pooled. Bootstrap analysis provided strong directional support for RMS, LF and HF power, and FR (**Figure S1**). MUAP amplitude showed greater uncertainty than the amplitude, spectral, and firing measures, whereas MUAP-duration estimates were comparable between muscles (median [IQR]: TA, 29.25 [29.08–29.48] ms; soleus, 29.03 [28.69–29.25] ms). Together, these features differed between healthy TA and soleus in directions consistent with their expected fast- and slow-twitch-like physiology.

**Figure 2.**
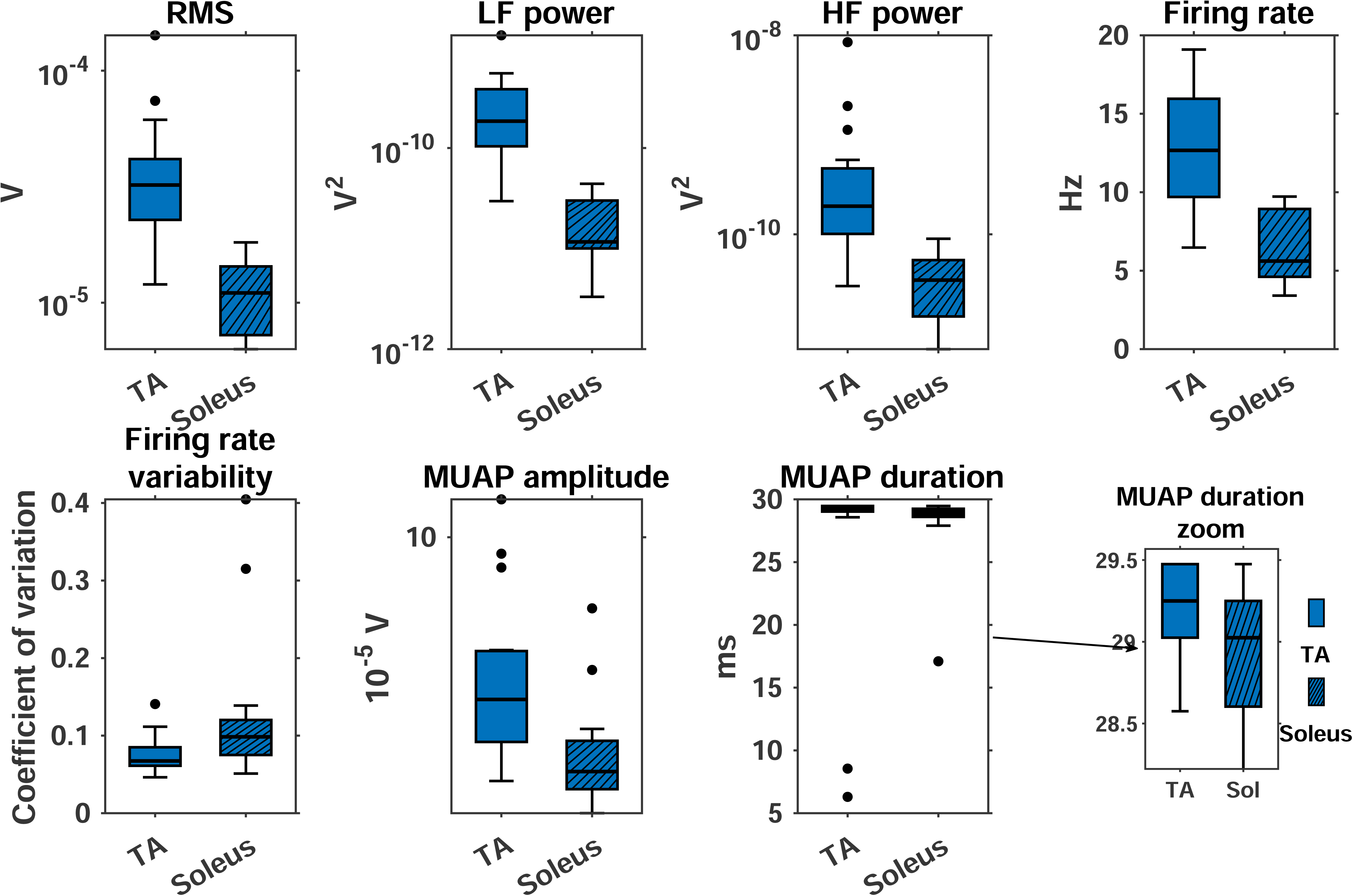
Healthy early-window TA-versus-soleus differences. Boxplots show early-window feature values for healthy TA and soleus contractions pooled across 30% and 50% MVC. RMS, LF power, HF power, and MUAP amplitude are displayed on logarithmic y-axes, whereas firing rate, firing rate variability, and MUAP duration are shown on linear scales. Individual points indicate values beyond the 1.5-IQR whiskers. The inset provides an expanded view of the main MUAP-duration distribution.

**Table 2.** Model-adjusted early-window HD-sEMG features and TA–soleus contrasts. Adjusted means were derived from maximum-likelihood linear mixed-effects models with Group, Muscle, MVC level, Group × Muscle, and a participant random intercept. Estimates were averaged equally across 30% and 50% MVC. Log-transformed features were back-transformed before reporting. Muscle comparisons represent Healthy TA versus soleus. Group comparisons represent ALS versus Healthy within TA. Residual degrees of freedom were used, and p-values were not adjusted for multiple comparisons. Model sample size was 51 observations for RMS, LF power, HF power, and MUAP amplitude and 49 for FR and FRV, across 17 participants.

| Feature | Healthy TA | Healthy soleus | TA vs. soleus | p (muscle) | p (Group within TA) | p (Group x Muscle) |
| --- | --- | --- | --- | --- | --- | --- |
| RMS ($\mu\text{V}$) | 33.8 | 10.3 | 3.29x | <0.001 | 0.478 | 0.875 |
| LF power ($\mu\text{V}^2$) | 204.3 | 14.0 | 14.55x | <0.001 | 0.640 | 0.645 |
| HF power ($\mu\text{V}^2$) | 239.1 | 31.3 | 7.65x | <0.001 | 0.0315 | 0.637 |
| FR (Hz) | 12.94 | 6.74 | +6.19 Hz | <0.001 | 0.382 | 0.667 |
| FRV (CV) | 0.0743 | 0.1313 | -0.057 | 0.022 | 0.133 | 0.192 |
| MUAP amplitude ($\mu\text{V}$) | 25.12 | 12.87 | 1.95x | 0.0010 | 0.911 | 0.755 |

### C. The TA-Versus-Soleus Profile Remains Detectable in ALS

Adjusted models confirmed higher RMS, LF and HF power, FR, and MUAP amplitude in healthy TA than soleus (all p≤ 0.0011; **Figure 3**; **Table 2**). Similar directional patterns were observed in ALS. FRV showed the opposite pattern, with lower adjusted values in healthy TA than soleus (0.074 vs. 0.131; p = 0.022). To characterize the ALS contrast directly, adjusted TA-to-soleus ratios were 3.39 for RMS, 16.76 for LF power, 6.23 for HF power, and 1.76 for MUAP amplitude. TA-minus-soleus differences were +5.42 Hz for FR and −0.004 for FRV. No Group × Muscle interactions were significant (all p ≥ 0.192), providing insufficient evidence that ALS altered the TA-versus-soleus contrasts, although nonsignificant interactions do not establish equivalence. Within TA, HF power was lower in ALS than in healthy controls (unadjusted p = 0.0315), whereas Group coefficients for the remaining features were nonsignificant (all p ≥ 0.133).

**Figure 3.**
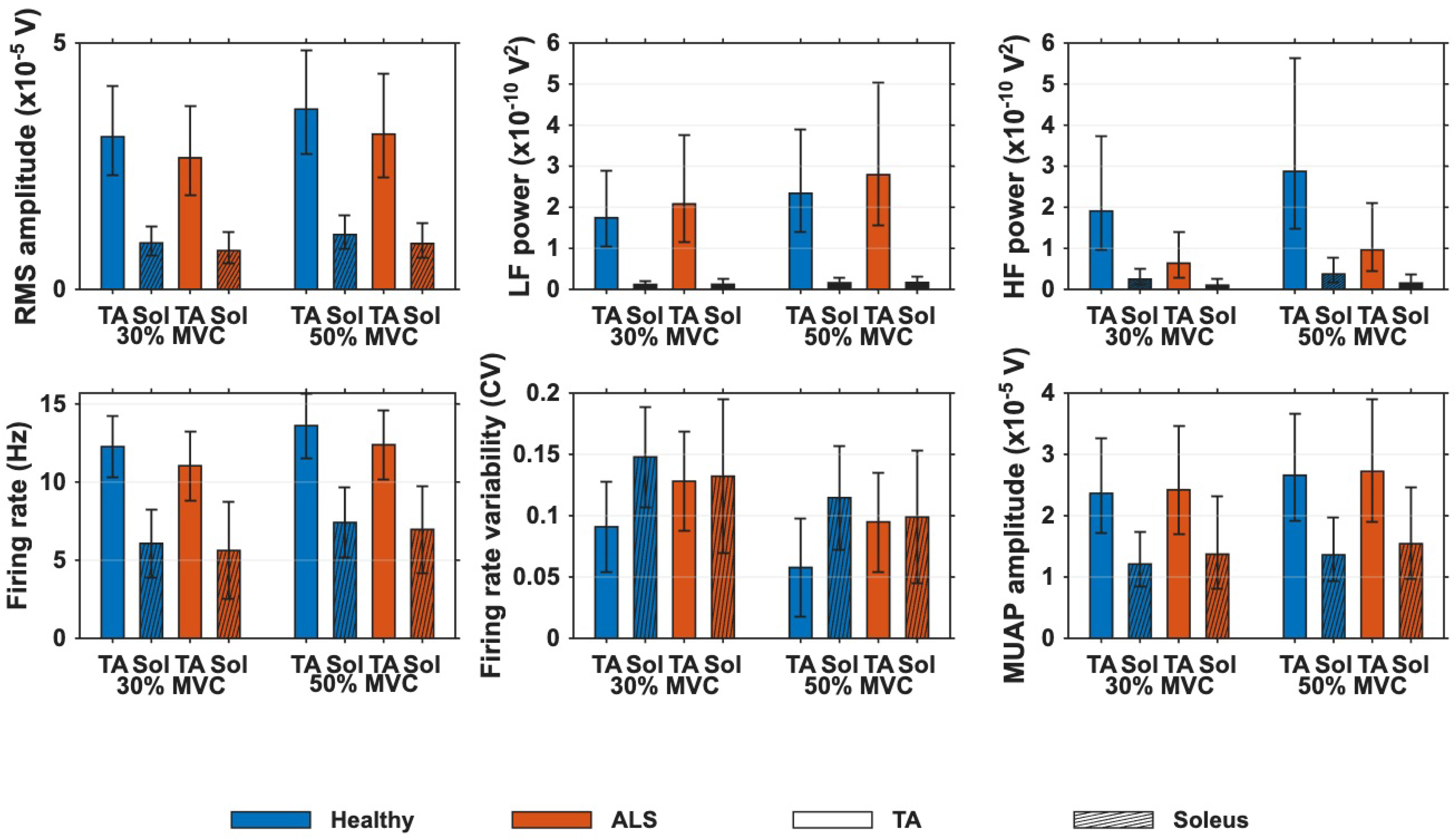
Adjusted early-window means in Healthy and ALS groups. Bars show model-adjusted means by group, muscle, and MVC level. RMS, LF power, HF power, and MUAP amplitude were modeled on the log scale and back-transformed for plotting. Error bars indicate 95% confidence intervals. This figure evaluates whether the TA-versus-soleus profile remains detectable in ALS during the early stable contraction window. No Group × Muscle interactions were significant. CV, coefficient of variation.

### D. Early-Versus-Late Ratios Suggest Greater TA Fatigue Sensitivity

Early-to-late fatigue ratios were examined descriptively and suggested larger declines in TA than soleus, particularly for RMS, HF power, and FR (**Figure 4**). Pooling observations across MVC levels, median changes in healthy TA were −15.0%, −45.6%, and −14.8% for these features, respectively, compared with +16.1%, +41.9%, and +21.8% in healthy soleus. In ALS, corresponding median changes were −30.0%, −64.2%, and −28.3% in TA versus +12.2%, +10.1%, and −1.1% in soleus. Increases were observed in 7 of 13 healthy soleus observations and 3 of 6 ALS soleus observations for each of the three features. However, fatigue ratios were more variable than early-window muscle contrasts. Because percentage change depends on the Early value, the same absolute change produces a larger ratio when the initial value is small.

**Figure 4.**
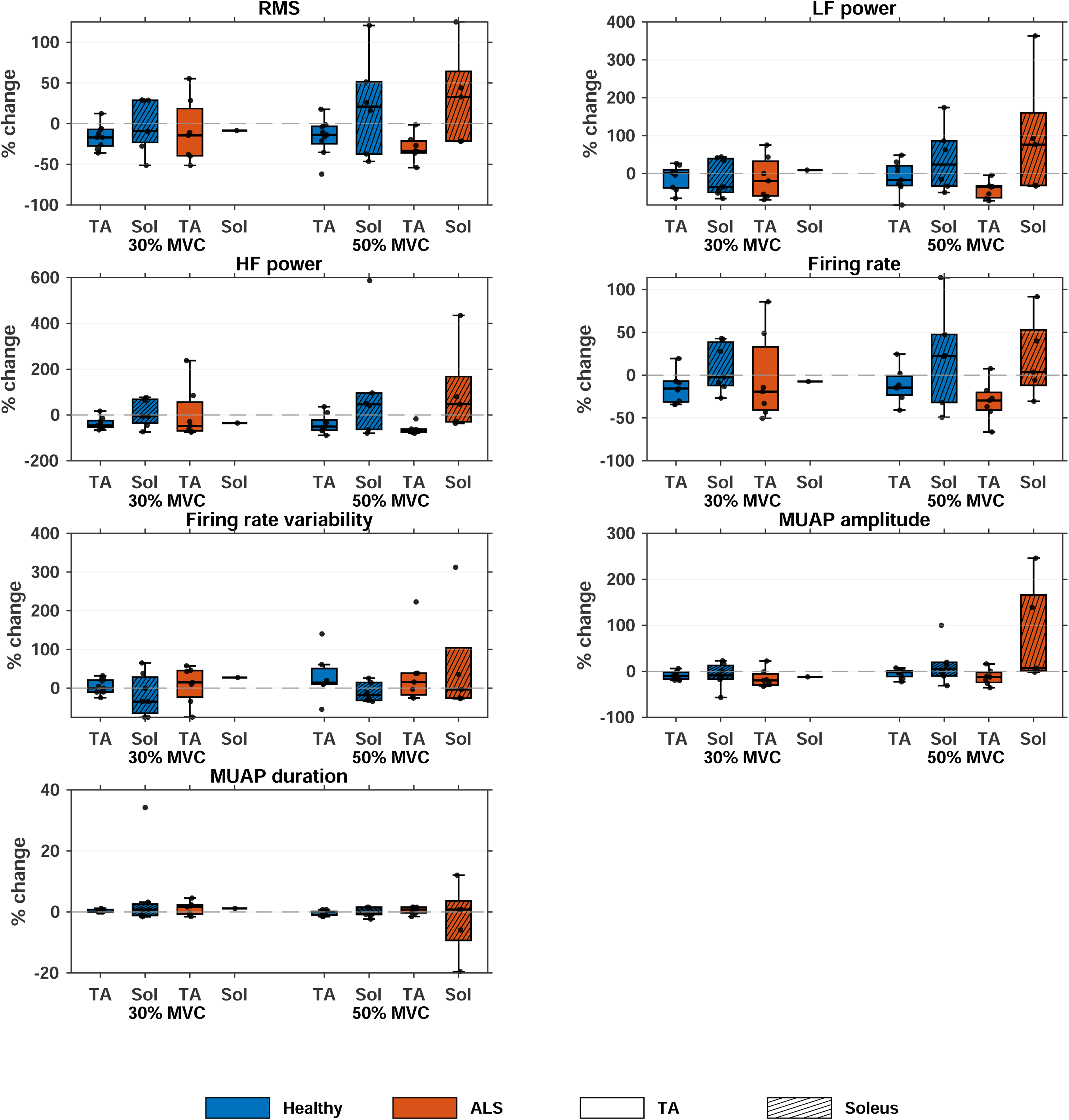
Early-versus-late fatigue ratios. Boxplots show percent change from the early to late analysis window by group, muscle, and MVC level. Negative values indicate a decrease over the sustained contraction. Y-axis ranges differ across panels to display the full distribution of each feature. At 30% MVC, ALS soleus data were available from only one participant; therefore, the corresponding orange soleus

This was particularly relevant to soleus, which often had lower Early values and fewer usable ALS observations. We therefore treated the fatigue ratios as supportive and placed greater emphasis on binned trajectories to characterize how features changed during sustained contraction.

### E. TA Shows a Stronger Time-Dependent Fatigue Trajectory

Across five time-normalized bins, TA showed greater descriptive declines than soleus, particularly for HF power, RMS, and FR (**Figure 5**). Median analyzed durations ranged from 145 to 295 s across group, muscle, and MVC combinations (**Table S2**). By Bin 5,median HF power had declined by 32.8% in healthy TA (n = 18 contractions) and 53.1% in ALS-affected TA (n = 14), compared with increases of 11.9% in healthy soleus (n = 13) and 41.5% in ALS soleus (n = 6). Corresponding median RMS changes were −7.6%, −19.9%, +4.2%, and +13.3%, respectively, while FR changes were −6.2%, −21.6%, +8.1%, and +30.2% (for FR, healthy TA n = 16; other sample sizes unchanged). These trajectories showed larger descriptive declines in ALS-affected TA than healthy TA. LF power was less consistent in healthy participants but showed a larger negative trajectory in ALS-affected TA than ALS soleus. MUAP amplitude changed in a similar direction, although less clearly than HF power, RMS, or FR. FRV and MUAP duration did not show distinct muscle-type fatigue patterns. Within TA, participants with ALS exhibited greater median final-bin declines in HF power, RMS, and FR than healthy controls, as quantified above. These between-group patterns were descriptive, and sparse ALS soleus observations limited assessment of whether ALS altered the TA-versus-soleus fatigue contrast.

**Figure 5.**
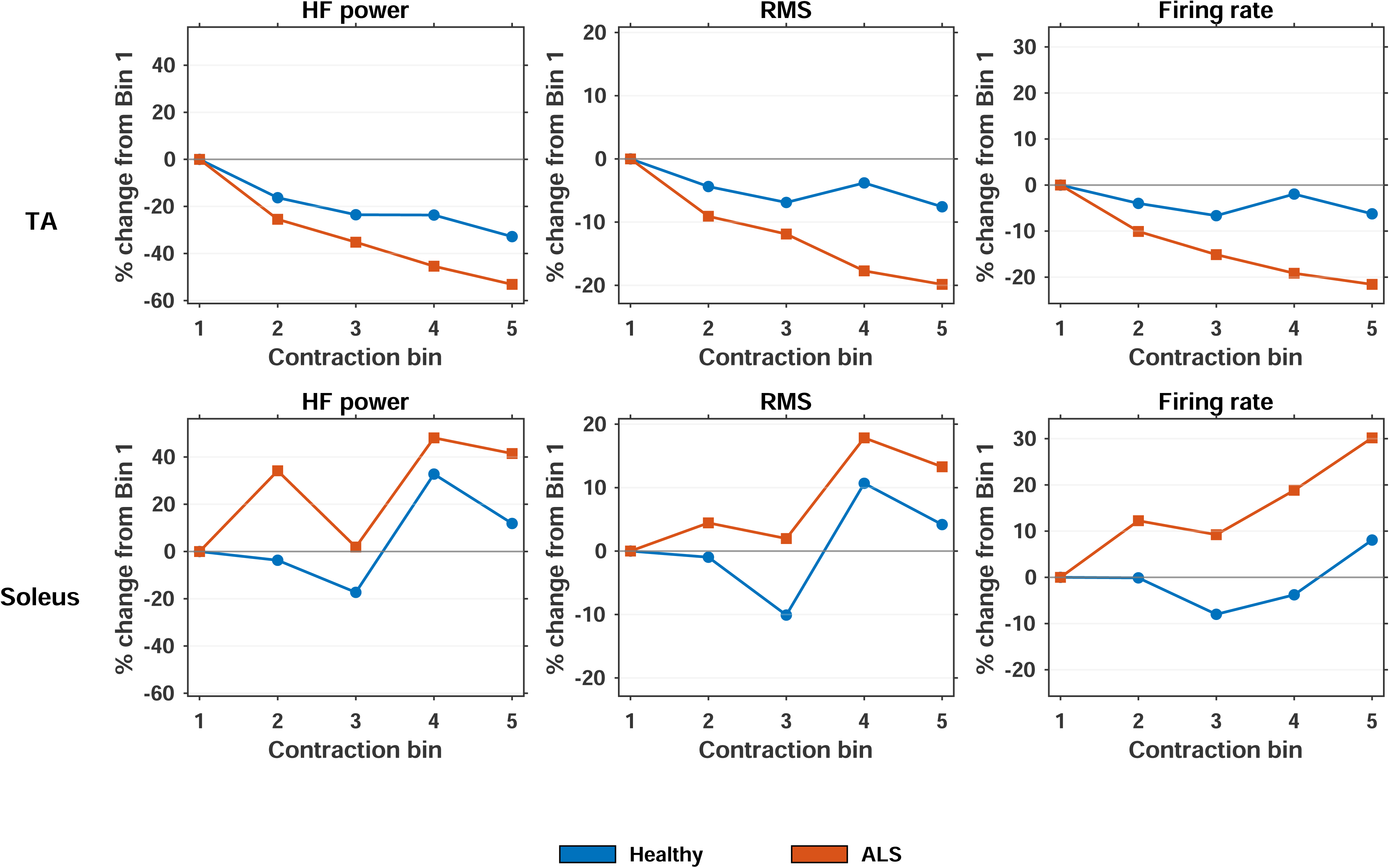
Binned fatigue trajectories. Lines show the median percent change from Bin 1 across five time-normalized contraction bins for HF power, RMS, and firing rate (FR), with 30% and 50% MVC contractions pooled. TA and soleus are displayed in separate rows, with Healthy and ALS trajectories shown within each panel. For each feature, TA and soleus panels use identical y-axis limits to preserve the relative magnitude of changes between muscles. TA generally showed progressive declines across bins, whereas soleus trajectories were more variable and, particularly in ALS, often increased later in the contraction.

### F. Post Hoc Robustness Analyses Support the Direction of the Binned Findings

Post hoc contraction-level slopes summarized the binned trajectories (**Figure 6**). HF power showed the strongest and most stable pattern: TA slopes were negative in both groups, whereas soleus slopes were generally positive or more variable. Leave-one-subject-out analyses preserved the main HF power directions. RMS slopes were similarly more negative in TA than soleus in both groups and more negative in ALS-affected TA than healthy TA. FR was directionally consistent with this pattern, although the healthy TA-versus-soleus direction was less stable. Direct comparisons of participant mean TA slopes between ALS and healthy controls did not show statistically significant differences for HF power (p = 0.438), RMS (p = 0.229), or FR (p = 0.238). LF power showed a more negative TA slope in participants with ALS than in controls, without a consistent healthy muscle contrast.

**Figure 6.**
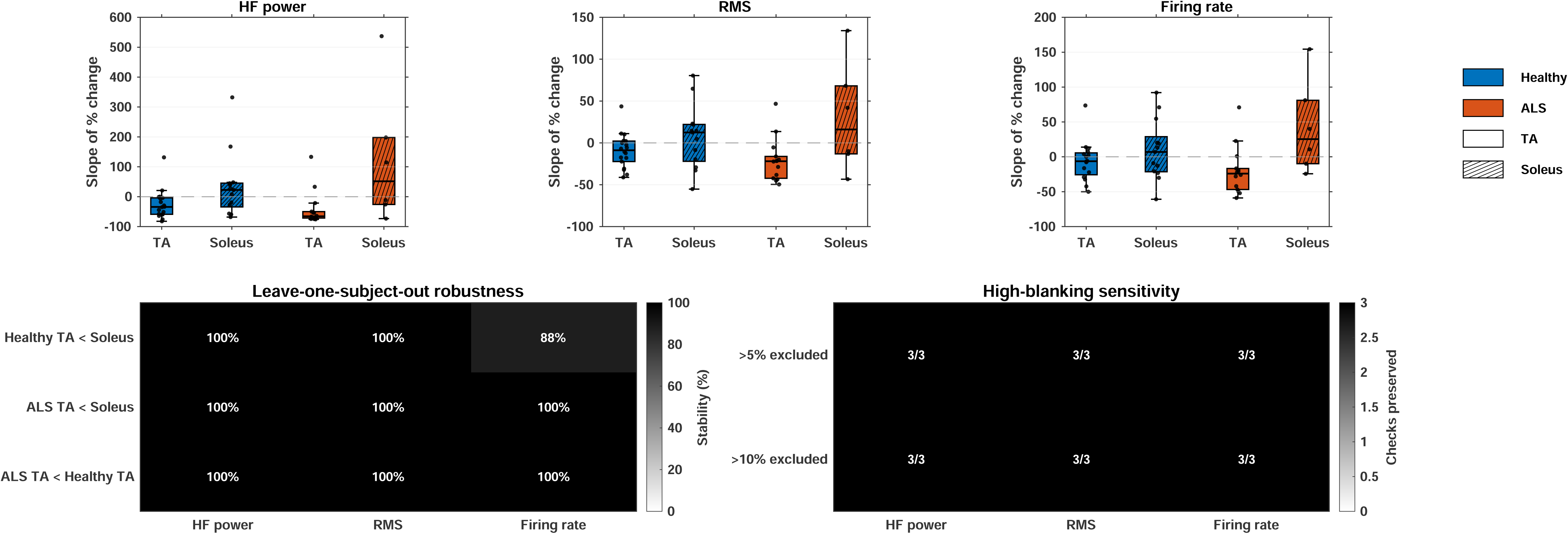
Binned fatigue slope and robustness summaries. Top panels show contraction-level slopes for percent change across bins. Y-axis ranges differ across the top panels to display the full distribution of slopes for each feature. Lower panels summarize leave-one-subject-out stability and high-blanking sensitivity for the main directional checks.

The main binned analysis retained contractions after conservative artifact blanking. Three contractions exceeded 5% blanking, including one healthy soleus contraction at 30% MVC and two ALS soleus contractions at 50% MVC, while none exceeded 10% (**Figure S2**). Excluding these three contractions left the principal HF power, RMS, and FR directions unchanged (**Figure 6**). Together with analyses that excluded each participant individually, these results suggest that the main trajectory patterns were not driven by individual participants or a small number of noisier recordings.

## 4. DISCUSSION

HD-sEMG distinguished TA from soleus through amplitude, spectral, motor unit firing, and MUAP features consistent with their contrasting physiology. The early-window profile persisted in ALS without significant Group × Muscle interactions. During sustained contractions, TA showed greater declines than soleus, particularly in HF power, RMS, and FR, with the largest descriptive declines in ALS-affected TA. These findings suggest that muscle physiology shapes HD-sEMG profiles in ALS and that fatigue responses warrant further investigation.

### A. Distinct HD-sEMG Profiles in TA and Soleus

The higher RMS, LF and HF power, FR, and MUAP amplitude in TA are consistent with its more phasic, relatively fast-twitch-like physiology compared with the postural, fatigue-resistant soleus. RMS and MUAP amplitude reflect several influences, including motor unit size, depth, territory, recording geometry, and activation (Henderson and McCombe, 2017; Lowery et al., 2004; Merletti and Farina, 2016). HF power may depend on action potential shape, conduction properties, and recruitment (Cifrek et al., 2009; Merletti et al., 2005; Merletti and Farina, 2016). Higher TA firing rates are also consistent with reports of generally lower discharge rates in soleus than TA (Inglis et al., 2025).

These features cannot identify fiber type directly. Architecture, depth, pennation, tissue thickness, and electrode placement influence surface EMG. TA is more superficial and may generate larger surface potentials independently of Type II fiber content, while soleus recordings may be attenuated by the overlying gastrocnemius muscle and tissue thickness (Henriksson-Larsén et al., 1983; Lowery et al., 2004; Merletti and Farina, 2016; Zwarts and Stegeman, 2003). Moreover, equal percentages of each muscle’s MVC do not ensure equivalent recruitment or discharge (Inglis et al., 2025). Handheld dynamometry may also underestimate plantarflexion strength, particularly when the examiner cannot fully resist stronger contractions (Marmon et al., 2013). Despite use of one examiner, this limitation could have altered relative MVC targets and fatigue responses. Accordingly, these measures represent muscle-specific electrophysiological profiles, not direct estimates of fiber composition. Although they cannot replace biopsy or imaging, their noninvasive and repeatable nature may offer potential for monitoring muscle activation, motor unit behavior, and fatigue-related changes over time, once test-retest reliability is established.

### B. The Early-Window TA-Versus-Soleus Profile Remains Detectable in ALS

TA remained higher than soleus for RMS, LF and HF power, FR, and MUAP amplitude in ALS. The adjusted HF power TA-to-soleus ratio was 6.23 (95% CI: 3.13-12.42), illustrating this contrast (**Figure 3**). No Group × Muscle interactions were significant, although this does not establish equivalence between groups. Within TA, HF power was lower in ALS than in healthy controls (unadjusted p = 0.0315), but the Group × Muscle interaction was not significant (p = 0.6365; **Table 2**). Thus, TA HF power differed by group without evidence of an ALS-specific change in the muscle contrast. Motor unit loss, altered recruitment, and reduced activation are possible contributors, but cannot be isolated here (Bashford et al., 2020; Henderson and McCombe, 2017; Nishikawa et al., 2022; Noto et al., 2023).

Incomplete soleus sampling substantially limits these comparisons. Of the five participants with ALS who contributed soleus data, none contributed at 30% MVC only, four contributed at 50% MVC only, and one contributed at both levels. This imbalance reduced the ability to detect interactions or determine whether ALS modifies muscle contrasts across contraction intensities.

### C. TA Shows Greater Time-Dependent Fatigue-Related Decline Than Soleus

Early-to-Late ratios suggested greater TA fatigue sensitivity. Binned trajectories showed more consistent declines in TA HF power, RMS, and FR, whereas soleus trajectories were flatter or more variable. ALS-affected TA had the largest median declines, but direct ALS-versus-healthy comparisons of TA slopes were nonsignificant for HF power, RMS, and FR (all p ≥ 0.229). HF power and FR were particularly informative because they distinguished the muscles early and showed prominent changes during sustained contractions. Sparse ALS soleus data nevertheless limited conclusions about whether ALS specifically altered the TA versus soleus fatigue contrast.

In SOD1 rodent models, fast-fatigable motor units and their neuromuscular junctions are preferentially vulnerable, with early loss reported in TA (Hegedus et al., 2008; Martineau et al., 2018; Pun et al., 2006). ALS models also show axonal loss, collateral sprouting, and reinnervation that may affect sustained motor unit activation (Martineau et al., 2018). Human HD-sEMG studies likewise indicate substantial remodeling, including faster characteristics among initially recruited motor units in ALS (Weddell et al., 2021). The larger TA declines could therefore involve reduced motor unit reserve, motor unit loss, altered recruitment, or compensatory reinnervation rather than fiber-type-related vulnerability alone. These mechanisms cannot be distinguished using the present observational electrophysiological measurements.

The principal fatigue features reflect overlapping mechanisms. Declining HF power may reflect altered conduction properties, action potential characteristics, or the active motor unit population (Cifrek et al., 2009; Enoka and Duchateau, 2008; Merletti et al., 2005; Merletti and Farina, 2016). Although RMS may initially increase through compensatory recruitment during sustained contractions, its decline may reflect reduced neural drive, fewer contributing motor units, changes in MUAP shape, or signal cancellation rather than motor unit loss alone (Dideriksen et al., 2011). Decreasing FR may indicate reduced discharge output or altered motor unit control (Enoka and Duchateau, 2008; Merletti et al., 2005; Merletti and Farina, 2016). Disease distribution, weakness, denervation, recruitment, self-reported fatigue, and task demands may also affect these trajectories (Martineau et al., 2018; Weddell et al., 2021). Therefore, these findings cannot establish preferential fatigue impairment of particular fiber types or fast-fatigable motor units.

Smaller or occasionally positive soleus changes may reflect greater fatigue resistance or different recruitment strategies, consistent with relative slow-unit resistance reported in ALS models (Pun et al., 2006). Percent-change metrics may also exaggerate soleus changes when Early values are small. However, animal motor unit findings do not translate directly to human whole-muscle HD-sEMG, and anatomy and sparse ALS soleus data complicate interpretation (Merletti and Farina, 2016; Zwarts and Stegeman, 2003). Flatter soleus trajectories should not be interpreted as absence of fatigue. Time normalization cannot fully account for differences in fatigue onset or task demands.

### D. Artifact Control and Robustness Checks

Artifact blanking reduced potential transient contamination of binned amplitude and spectral estimates. Only three contractions exceeded 5% blanking: one healthy soleus contraction at 30% MVC and two ALS soleus contractions at 50% MVC. None exceeded 10% (**Figure S2**). Because all three involved soleus, this exclusion check was limited. Furthermore, artifact blanking could inadvertently remove large physiological MUAPs or fasciculation potentials, which may occur more frequently in ALS (Tamborska et al., 2020). The main HF power, RMS, and FR trajectory directions persisted in analyses that excluded each participant individually and after excluding the three contractions. These checks support directional stability.

### E. Future Directions

Larger, balanced cohorts with complete soleus recordings at both force levels are needed to test Group × Muscle effects and distinguish muscle-specific from disease-related changes. Longitudinal studies should assess aging and ALS progression alongside test-retest reliability, including during fatigue. Test-retest reliability of these features, including during fatigue, should also be established. Additional force levels with sufficient rest would address contraction-intensity effects and residual fatigue.

Ultrasound could quantify anatomical influences such as muscle depth, pennation, and tissue composition (Barnes and Simon, 2019; Pillen and van Alfen, 2011). Validation against biopsy, imaging, or independent muscle-composition measures remains necessary before interpreting HD-sEMG features as direct fiber-type indicators.

## 5. CONCLUSION

HD-sEMG distinguished TA from soleus in healthy participants using amplitude, spectral, FR, and MUAP amplitude features. Their directions were consistent with fast- and slow-twitch-like physiology, supporting noninvasive assessment of muscle-type-related electrophysiological profiles. In ALS, the early-window TA-versus-soleus profile remained detectable, and no early-window Group × Muscle interaction was significant. Exploratory fatigue analyses showed greater declines in HF power, RMS, and FR in TA than soleus. These declines were largest in ALS-affected TA, although differences in TA fatigue slopes between groups were not statistically significant. These findings warrant validation in larger, balanced cohorts.

## Author Contributions

Ernesto Bedoy: Study design; Methodology (development of the HD-sEMG analytical framework, feature selection, signal preprocessing, motor unit decomposition strategy, fatigue-trajectory analysis, and statistical modeling); Investigation (study piloting, experimental execution, and protocol implementation); Data acquisition (HD-sEMG recordings); Software (development of MATLAB analysis pipeline); Data curation; Formal analysis (motor unit decomposition, feature extraction, and statistical analysis); Visualization; Writing - original draft. Maia Brown: Investigation (protocol approval, study piloting, participant recruitment, and dynamometer application).

Breanna Sullivan: Investigation (participant recruitment, informed consent, dynamometer application, and data acquisition support); Clinical coordination.

Michael Christofidis: Investigation (study piloting, study-session support).

Tawfiq Al-lahham: Clinical expertise; Methodology (clinical interpretation of EMG); Investigation (clinical demonstration of needle EMG and interpretation of findings; dynamometer training); Resources (clinical space coordination).

Douglas Weber: Conceptualization; Supervision.

Christi Kolarcik: Conceptualization; Funding acquisition; Study Design; Supervision; Investigation (protocol approval and implementation, participant recruitment, informed consent, and study piloting); Clinical coordination.

*All authors reviewed, edited, and approved the final manuscript*.

## Abbreviations

ALS: amyotrophic lateral sclerosis
EMG: electromyography
FR: firing rate
FRV: firing rate variability
HD-sEMG: high-density surface electromyography
HF: high frequency
IQR: interquartile range
LF: low frequency
MUAP: motor unit action potential
MVC: maximum voluntary contraction
RMS: root mean square
SNR: signal-to-noise ratio
TA: tibialis anterior.

## Data Availability

All data produced in the present study are available upon reasonable request to the authors.

The data that support the findings of this study are available from the corresponding author upon reasonable request, subject to institutional and ethical restrictions. Analysis code and experimental protocols will be made publicly available upon acceptance.

## Acknowledgments

This work was supported by The ALS Association (Grant No. 24-SGP-723). The authors wish to thank all study participants for their time and participation. We also acknowledge the clinical staff at the University of Pittsburgh Medical Center (UPMC) for assistance coordinating clinical space for data collection. Finally, the authors are grateful to Brandy Norwood (*Brandy*) for inspiring the completion of this work done in memory of Blase Urban.

---

**Figure S1.**
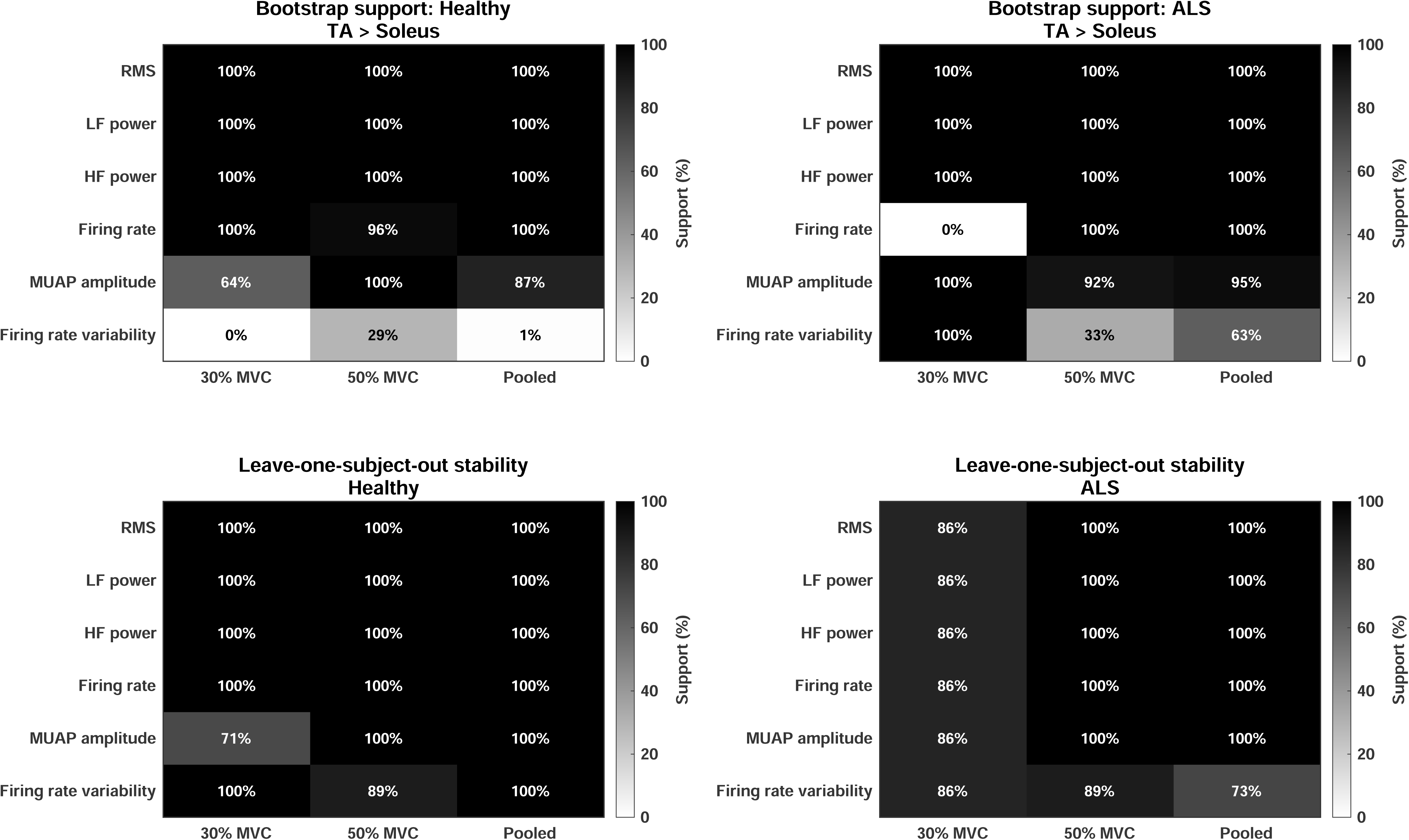
Feature-specific bootstrap and leave-one-subject-out summaries for baseline TA-minus-soleus contrasts

**Figure S2.**
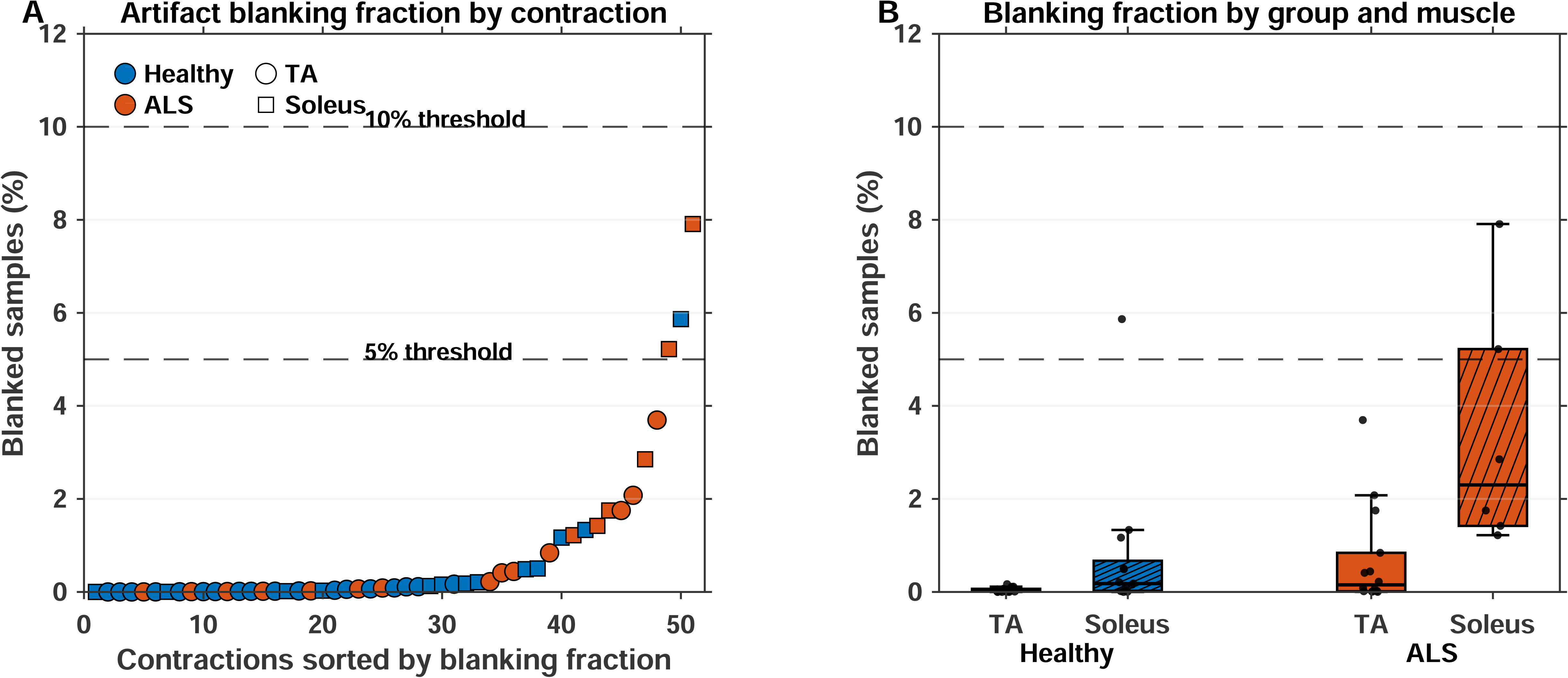
Artifact blanking summary

