## Supplementary Information for "High-Density Surface Electromyography Detects Fiber-Type-Related Profiles and Disease-Relevant Fatigue Trajectories in Amyotrophic Lateral Sclerosis"

#### Detailed HD-sEMG Acquisition and Decomposition

Before electrode placement, the skin was cleaned and lightly abraded with Lemon Prep abrasive gel (Mavidon, Flat Rock, NC) to improve signal quality and reduce impedance. HD-sEMG was collected using a Trigno Galileo decomposition sensor (Delsys Inc., Natick, MA, USA). The sensor contains four 99.99% silver detection contacts arranged with 5-mm inter-electrode spacing. For TA recordings, the reference sensor was placed on the soleus. For soleus recordings, the reference sensor was placed on the TA. EMG signals were differentially amplified and digitized at the sensor with 16-bit resolution and a sampling rate of 2222 Hz. Motor unit spike trains were obtained using NeuroMap software (v1.2.2, Delsys Inc.), which uses the Precision Decomposition III algorithm (De Luca et al., 2015, 2006; Nawab et al., 2010).

#### Signal Processing and Motor Unit Quality Control

Raw HD-sEMG recordings were screened before MATLAB analysis to exclude recordings with poor signal quality. Recordings were excluded if the SNR was less than 3 dB. SNR was calculated as 10 log_10_ (P_contraction_/ P_baseline_), where P_contraction_ was the EMG power during contraction and P_baseline_ was the baseline noise power. Motor unit decomposition accuracy was evaluated using the Decompose-Synthesize-Decompose-Compare method (De Luca et al., 2015, 2006; Nawab et al., 2010). For the present analysis, only motor units with decomposition accuracy ≥90% were retained before feature extraction. Additional prespecified physiological plausibility filters excluded motor units with peak FRs greater than 60 Hz, mean FRs less than 2 Hz, or inter-pulse interval coefficient of variation greater than 1.0. These study-specific quality-control criteria were used to remove units with implausibly high discharge rates, sparse firing, or highly irregular firing behavior.

For early-versus-late MUAP features, a motor unit was included only if it had at least 20 spikes in both the Early and Late analysis windows. For the binned MUAP analysis, the threshold was set to 10 spikes per bin because each bin represented a shorter segment of the contraction than the 10-second Early and Late windows. These thresholds were analysis quality-control criteria used to avoid estimating MUAP morphology from sparse spike-triggered averages. Spike-triggered average snippets containing blanked artifact samples were excluded.

#### MUAP Waveform and Duration Calculation

MUAP waveforms were estimated using spike-triggered averaging with a 30-ms total window around each motor unit discharge. MUAP duration was calculated separately for each channel as the time between the first and last samples at which the absolute waveform exceeded 10% of its maximum absolute amplitude. Channel-level durations were summarized using the median for each motor unit, followed by the median across retained motor units. Early and Late MUAP durations were calculated separately from their respective analysis windows.

#### Feature-Specific Data Availability

The dataset was unbalanced because soleus recordings were less complete than TA recordings, particularly in ALS (**Table S1**). Two TA 50% MVC trials contributed raw EMG amplitude and spectral features to the binned analysis but did not contribute FR or MUAP features because bin-level motor unit information did not meet quality-control criteria.

#### Binned Fatigue Normalization

For each Subject × Muscle × MVC observation, bin values were normalized to Bin 1:

*Normalized bin value = Bin value / Bin 1 value.*

Percent change from Bin 1 was calculated as:

*Percent change from Bin 1 = 100 × (Bin value − Bin 1 value) / Bin 1 value.*

#### Spike Artifact Blanking and Sensitivity Analysis

Isolated spike artifacts can distort RMS and spectral estimates, especially in binned analyses (Cifrek et al., 2009; Merletti and Farina, 2016). A conservative artifact-blanking procedure was therefore applied to filtered EMG before bin-level feature extraction. The goal was to remove brief nonphysiological transients while preserving true high-force EMG activity.

Artifact thresholds were estimated separately for each channel using the stable contraction period. The channel median and median absolute deviation were calculated from the stable contraction window, and the median absolute deviation was converted to an estimate of signal variability (Leys et al., 2013). Samples were labeled as artifact candidates only when they exceeded a high threshold relative to stable-window variability; a multiplier of 12 times the estimated standard deviation was used. A region extending 8 ms on each side of each detected artifact was blanked and excluded from RMS and spectral calculations. MUAP spike-triggered average snippets containing blanked EMG were also excluded. The 8-ms padding window was selected to remove brief transients and immediate edge effects.

No contraction was excluded from the main binned analysis based only on blanking fraction. Instead, the percentage of blanked samples was logged for each contraction. Post hoc sensitivity analyses repeated the trajectory analysis after excluding trials with blanking fractions greater than 5% and 10% to determine whether the main fatigue trajectories depended on higher-blanking trials. Three contractions exceeded the 5% blanking threshold, whereas none exceeded 10%; therefore, the 10% sensitivity analysis retained the full dataset.

### **Supplementary Table**

| **Group** | **Total Participants** | **TA 30%** | **TA 50%** | **TA both MVCs** | **Soleus 30%** | **Soleus 50%** | **Soleus both MVCs** | **Complete TA/Sol at both MVCs** |
| --- | --- | --- | --- | --- | --- | --- | --- | --- |
| Healthy | 10 | 9 | 9 | 9 | 7 | 6 | 6 | 5 |
| ALS | 7 | 7 | 7 | 7 | 1 | 5 | 1 | 1 |
| **Table S1. Usable recording availability by group, muscle, and MVC level.** Values indicate the number of participants contributing usable observations to each analysis set. | | | | | | | | |

### **Supplementary Figures**

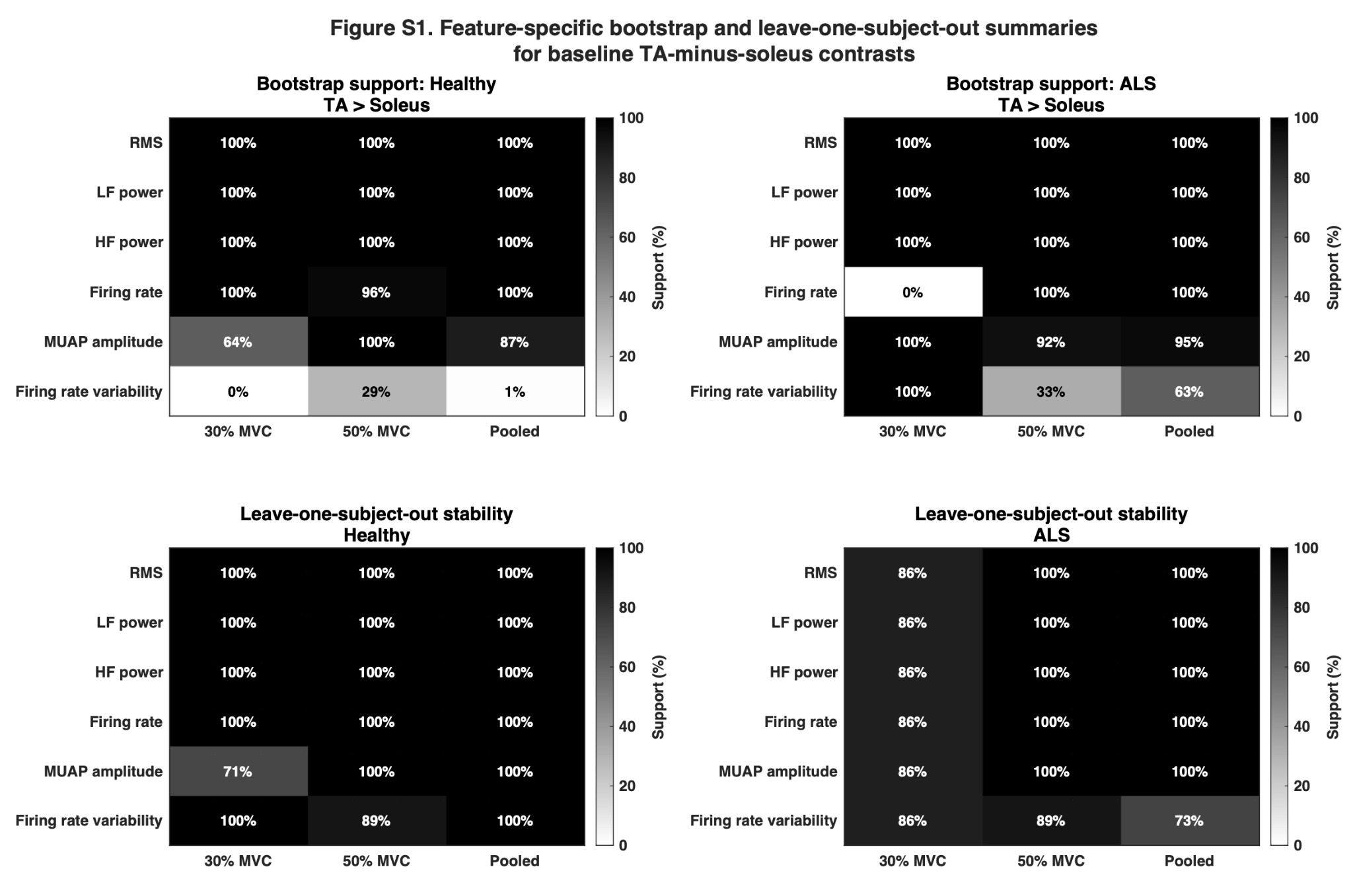

**Figure S1. Early-window TA-versus-soleus robustness summaries.** Heatmaps show bootstrap support and leave-one-subject-out stability for the direction of early-window TA-minus-soleus contrasts.

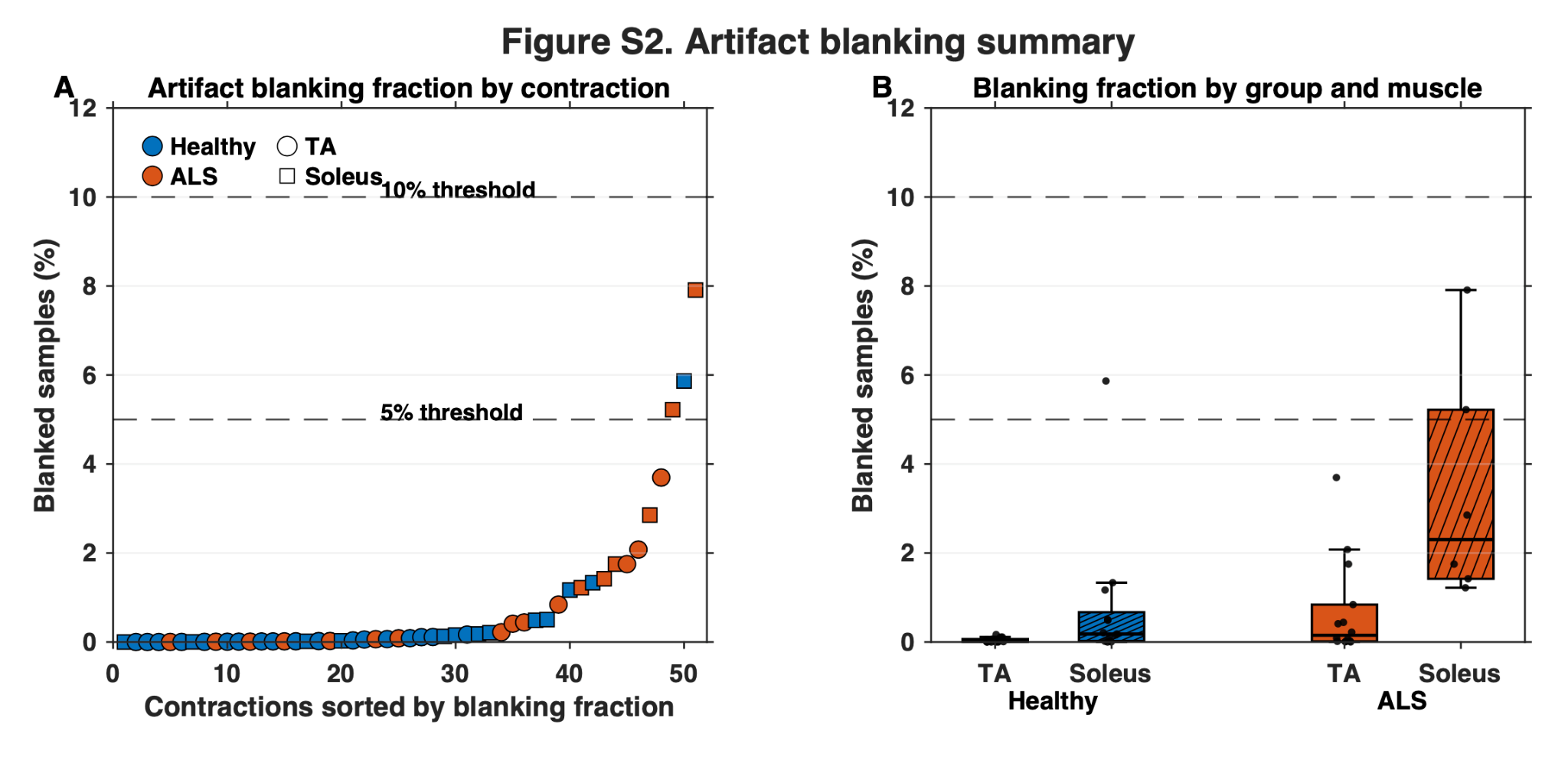

**Figure S2. Artifact blanking summary.** Panel A shows the percentage of blanked samples for each contraction, sorted by blanking fraction. Panel B summarizes blanking fractions by group and muscle. Dashed lines mark the 5% and 10% thresholds used for high-blanking sensitivity analyses.
